# Long term vaccination strategies to mitigate the impact of SARS-CoV-2 transmission: a modelling study

**DOI:** 10.1101/2023.02.09.23285743

**Authors:** Alexandra B Hogan, Sean L Wu, Jaspreet Toor, Daniela Olivera Mesa, Patrick Doohan, Oliver J Watson, Peter Winskill, Giovanni Charles, Gregory Barnsley, Eleanor M Riley, David S Khoury, Neil M Ferguson, Azra C Ghani

**Author notes:** Correspondence: Dr Alexandra Hogan, Professor Azra Ghani.

## Abstract

**Background:** Vaccines have reduced severe disease and death from COVID-19. However, with evidence of waning efficacy coupled with continued evolution of the virus, health programmes need to evaluate the requirement for regular booster doses, considering their impact and cost-effectiveness in the face of ongoing transmission and substantial infection-induced immunity.

**Methods and findings:** We developed a combined immunological-transmission model parameterised with data on transmissibility, severity, and vaccine effectiveness. We simulated SARS-CoV-2 transmission and vaccine rollout in characteristic global settings with different population age-structures, contact patterns, health system capacities, prior transmission, and vaccine uptake. We quantified the impact of future vaccine booster dose strategies with both original and variant-adapted vaccine products, in the presence of both continuing transmission of Omicron subvariants and considering the potential future emergence of new variants with modified transmission, immune escape, and severity properties. We found that regular boosting of the oldest age group (75+) is the most efficient strategy, although large numbers of hospitalisations and deaths can be averted by extending vaccination to younger age groups. In countries with low vaccine coverage and high infection-derived immunity, boosting older at-risk groups is more effective than continuing primary vaccination into younger ages. These findings hold if even if virus drift results in a gradual reduction in vaccine effectiveness over time due to immune escape. In a worst-case scenario where a new variant emerges that is 10% more transmissible, as severe as Delta, and exhibits substantial further immune escape, demand on health services could be similar to that experienced during 2020.

**Conclusions:** Regular boosting of the high-risk population remains an important tool to reduce morbidity and mortality from current and future SARS-CoV-2 variants. The cost-effectiveness of boosting is difficult to assess given the ongoing uncertainty in the likelihood of future variants and their properties but focusing vaccination in the highest-risk cohorts remains the most efficient strategy to reduce morbidity and mortality.

## Introduction

The rapid development and delivery of vaccines to protect against COVID-19 infection and disease dramatically altered the course of the pandemic, saving an estimated 19.8 million lives in the first year of vaccination alone.^1^ However, the effectiveness of COVID-19 vaccines wanes, with considerable declines against infection but slower declines against severe disease and death. Thus, it is likely that continued booster programmes will be needed to maintain high effectiveness against severe disease and death, particularly in those at highest risk of more severe outcomes. In addition, vaccine booster programmes have been successful in partially restoring effectiveness against severe disease and death when levels of existing vaccine protection have been eroded by the emergence of new variants that have resulted in immunological escape.^2–4^

Many countries are now considering how best to schedule regular boosting to protect against ongoing endemic circulation of the virus as well as against future epidemic waves with Omicron subtypes or new variants. The benefit of such strategies in any given population will depend on the current stage of their vaccine programme, including the supply of vaccine doses and the extent that these doses are matched to the current circulating strains, as well as the magnitude of the epidemic that has been experienced to date and thus the extent of infection-acquired immunity. Furthermore, the benefits of booster vaccination will depend on the extent to which any future variant replaces the current Omicron variant, and whether it further evades existing immunity.

Throughout the COVID-19 pandemic, mathematical and computational modelling has been a key component of longer-term planning and has been widely used to inform decisions on future vaccine strategies.^5^ A number of studies have focussed on country-specific projections of epidemic progression and vaccine impact, or have focussed on allocation or prioritisation of the limited supply of doses (particularly in the early stages of vaccine rollout).^6–8^ More recently, models have been developed to consider longer-term strategies for continued vaccination using assumed profiles for vaccine-induced and infection-induced immunity over time.^9–11^ Planning for these scenarios, particularly considering potential future variant characteristics, has been identified as a global public health priority.^12^

In contrast to other modelling studies, we sought to understand patterns of COVID-19 disease dynamics in the context of hybrid immunity (immunity induced by both infection and vaccines), in order to capture the interactions between past exposure and vaccination. We did this by applying an existing within-host model of underlying immunity dynamics and protection against infection and severe disease (similar to that presented in Khoury et al^13^), that has been previously fitted to vaccine effectiveness data from England^14^ and embed this model within a population-based virus transmission model for SARS-CoV-2. We then use this model to explore timelines for transition to endemic circulation, and the impact of different targeted booster strategies including the use of the bivalent vaccines. We additionally consider the potential impact of the gradual emergence of new variants and the likelihood that vaccination could sufficiently mitigate their impact.

## Methods

### Immunological model

The immunological model is as described in Hogan *et al*.^14^ Briefly, we followed the approach in Khoury *et al*, in which neutralizing antibody titre is assumed to be a correlate of protection against SARS-CoV-2 infection, severe disease, and death over time. Such a model does not necessarily exclude other immune mechanisms playing a role in protection – including T-cell mediated immunity – but rather makes the underlying assumption that the patterns of protection over time can be related to the trends observed in the neutralising antibody titre. We therefore define these underlying dynamics of immunity as an individual’s immunity level (IL).^14^ We further assume that an individual’s IL decays according to a biphasic exponential decay function, where an initial faster period of decay is followed by a longer period of slow decay.^13^ We then assume logistic relationships between IL and effectiveness to capture time-varying vaccine protection against mild disease (infection) and hospitalisation over time, with the logistic function parameterisation capturing higher protection against severe outcomes.^13^ The model parameters against the Delta and Omicron variants for three vaccine products – the Oxford/AstraZeneca AZD1222 vaccine, the Pfizer-BioNTech BNT162b2 vaccine and the Moderna mRNA-1273 vaccine – in combinations for the primary series and boosting products are reproduced in **Table S1**. To capture loss of immune recognition against Omicron and future variants, we estimate a multiplicative scaling factor (referred to as the variant fold reduction, VFR) to reflect the reduced neutralization of a given variant for all vaccines modelled. This VFR for the original vaccines is based on our estimates of the degree of immune escape obtained by fitting to vaccine effectiveness data against the Delta and Omicron (BA.1/2) variants.^14^ We then reduce these estimated VFRs for the Moderna mRNA-1273.214 bivalent vaccine based on immunological data from recent trials^15^ as described in Hogan et al.^14^

We use the same approach to capture infection-induced immunity and its interaction with vaccine-induced immunity, where each infection is assumed to generate a boost to IL of 1, i.e. equivalent to that measured in convalescents,^13^ which corresponds to a mean protection against re-infection of 67% over 180 days, at the lower end of estimates obtained in a recent study of infection-induced protection against Omicron BA.4 and BA.5 subvariants which found 76.2% (95% CI: 66.4%-83.1%) protection against symptomatic re-infection.^16^ This level of boost is higher than that observed under vaccination, potentially representing a broader and longer-lasting immune response, and resulting in higher infection-induced protection. We additionally performed sensitivity analyses to this assumption exploring boosts to IL of 0.75 and 1.25, corresponding to protection against re-infection over 180 days of 50% and 81% respectively. The infection-induced IL is assumed to decay at the same rate as following booster vaccination. Each infection or vaccine dose results in an additive increase in IL, with an upper limit on the total level of vaccine- or infection-induced IL for each individual.^17^

We assume that following the emergence of a variant, the infection-induced IL developed through exposure to previous variants is reduced by the VFR in the same way that vaccine-induced protection is reduced; but that the infection-induced IL developed to the new variant is not reduced. This captures strain-specific protection against infection without explicitly modelling each variant (see Supplementary Material).

### Population model and vaccine allocation

To explore the population impact of vaccines, we developed a stochastic, individual-based model of SARS-CoV-2 transmission and vaccination (open-source at https://mrc-ide.github.io/safir/).^18^ Vaccine-derived and infection-induced immune dynamics follow the immunological model described above. The model structure and epidemiology broadly mirror a previously published compartmental model, but these processes are instead implemented at the individual level.^19^ This allows for immunity to be implemented at the individual level, capturing both vaccine- and infection-induced immunity, individual variation in this immunity, decay over time, and allowing for individual-level tracking of vaccine and infection history. It also allows for a high degree of flexibility in dose and age-based vaccine prioritisation strategies. Transitions between epidemiological states are summarised in **Figure S1** and **Table S2**, with natural history parameters for SARS-CoV-2 infection, age-stratified probabilities of requiring hospital care and the infection fatality ratio as in Hogan *et al* (**Table S3**).^14^ The model captures differences between countries in demography, age-mixing patterns, and access to hospital facilities.^19,20^

Vaccines are allocated according to an algorithm accounting for available stock, the age groups that are prioritised for each dose, minimum time delays between the receipt of subsequent doses, and coverage targets for each dose and age group (**Supplementary Material S1.5**; **Figure S3**).^19^

### Settings and transmission

We consider two representative income settings – high-income countries (HIC) and lower-middle-income countries (LMIC) – and characterise each setting by contact patterns and demography.^19,20^ In LMICs we assume healthcare system capacity is limited; once modelled hospitalised cases exceed a threshold in these settings, infected individuals who require hospital care experience worse outcomes.^20^ In HICs we assume no limit to healthcare capacity due to surge provisions.

We further stratify the current epidemiological state of countries into three categories. “Category 1” represents countries that have experienced substantial past transmission (and hence have a substantial level of infection-induced immunity) alongside a high level of access to vaccines. Many high- and upper-middle-income countries fall into this category – including countries in North America, Central/South America, the Middle East and Europe. We created a representative epidemic profile for such countries, with a first wave occurring between March and May 2020, a second wave during the northern hemisphere winter of 2020/21, and transmission gradually increasing (interventions being relaxed) from mid-2021 (**Figure S2**). This broadly characterises a northern hemisphere setting but does not include seasonality. “Category 2” are countries that have experienced substantial prior transmission and have had limited distribution of vaccines. Many low- and lower-middle-income countries fall into this category (although we note that several LMICs have successfully limited transmission). For these countries we model a similar background epidemic to that in Category 1 (**Figure S2**) but with fewer interventions in place during 2021. “Category 3” countries are those that successfully interrupted transmission for a substantial time period (“zero-COVID” countries, mostly in east Asia and the Pacific) and therefore have more limited infection-induced immunity, alongside high vaccine uptake. For these we assume a gradual lifting of restrictions (**Figure S2**) from late 2021.

In all settings we assume that the Omicron variant (BA.1 and BA.2 subtypes) gradually replaces Delta over one month from end-November 2021. This replacement impacts infection- and vaccine-induced immunity, transmissibility, and severity (see **Supplementary Material**). We did not explicitly model the impact of the BA.4/BA.5 or subsequent Omicron subtypes; however, these are implicitly captured in the scenarios in which a gradual drift is assumed (see Variant Scenarios).

### Vaccine dose strategies

For all scenarios we prioritise the oldest individuals with vaccines delivered sequentially to consecutive 5-year age groups until the target age group is vaccinated. We assume total maximum population-level coverage of 80% and 53% for the primary series and booster doses respectively in HIC settings, based on World Health Organization reported coverage by income setting.^21^ In LMIC settings we assume a maximum of 52% and 34% for the primary series and booster doses respectively (compared to the reported population-level uptake of 55% and 8% on 18 July 2022).^21^ In all settings, uptake is assumed to be highest in older age groups.^22,23^ We consider an additional scenario where 70% population-level coverage of the primary series is achieved in LMIC settings, based on World Health Organization policy targets.^24^ As we assume that only individuals 10 years and older (10+) are eligible for vaccination, this total population-level coverage corresponds to higher uptake within targeted groups and zero coverage within ineligible groups; the within-age group uptake for each setting is shown in **Table S4**.

Vaccine distribution strategies are implemented as follows. For Categories 1 and 3 (HICs), vaccines are administered at a constant rate of 5% of the population receiving one dose per week, starting 1 January 2021, assuming the mRNA-1273 vaccine for the primary 2-dose series and booster to those 10+. After the primary doses and first booster dose, we then either cease to administer any additional doses; administer either annual or 6-monthly booster doses of the mRNA-1273.214 bivalent vaccine to the 75+ population at the same pace; administer these same schedules to the 60+ population with mRNA-1273.214; or boost the 10+ population annually with mRNA-1273.214. We additionally consider the outcome if all doses from dose 4 onwards are the original mRNA-1273 vaccine instead of the bivalent product.

For Category 2 (LMICs), vaccines are administered at a maximum constant rate of 2% of the population immunised per week with starting 1 April 2021. We assume the first two doses are the AZD1222 vaccine, the first booster (third dose) is with the mRNA-1273vaccine, and subsequent doses are either with the bivalent mRNA-1273.214 (default) or the original mRNA-1273 vaccine. We assume the primary series and first booster is administered to those aged 10+ according to the levels of uptake in **Table S4**. We then either cease to administer any additional doses; administer annual booster doses to the 60+ population at the same pace; boost the 40+ population annually; or boost the 10+ population annually.

For Category 2 (LMICs), we additionally model a separate scenario where we consider the relative impact of administering doses to vaccinate the younger working-age population with their primary series, versus diverting those doses to vaccinate the older population with a booster dose. We commence vaccination with the AZD1222vaccine from April 2021, delivering the primary 2-dose series to the 40+ population. Once the target coverage is achieved, vaccination is paused until the delay between the second and booster doses (12 months) is complete. We then construct the following scenarios. For the first, we vaccinate the 40+ population with booster doses, beginning with the oldest (80+) age group. For the second, we take the same number of doses that would be required to give boosters to 40+ years, and instead deliver these doses to individuals younger than 40 years (2 doses per person). We construct these rollout scenarios such that the daily doses delivered is equivalent between scenarios. We compare these outputs with the scenario where no additional doses are delivered beyond 2 doses to the 40+ population.

### Variant scenarios

For the main analysis, we first model a scenario in which no additional variants emerge beyond the initial Omicron variant. We then additionally consider two possibilities for variant emergence. In the first, we consider a situation where the virus continues to evolve or “drift”, with a new variant regularly replacing the dominant variant. This can be considered to represent the gradual drift that is now being observed with the Omicron sub-variants. We implement this in the model by increasing both the level of transmission and the VFR (relative to Delta) every 4 months, to represent both a small increase in transmissibility and gradual immune escape. Given that the frequency and magnitude of the future viral evolution of SARS-CoV-2 is challenging to predict, we consider two illustrative levels of increase for both transmission and immune escape, of 5% and 10%, referred to as “5% drift” and “10% drift” respectively.

For the second type of variant scenario, we simulate the emergence of a single new dominant variant between 1–31 October 2023 and assume that the new variant remains dominant for the remainder of the simulation time period (i.e. to end-2024). We consider three possible new variants with the following characteristics: (1) severity increased to that of Delta (“increased severity”); (2) VFR relative to Delta increased to 10 (“additional immune escape”) and hence an additional 2-fold reduction relative to Omicron (an antigenic shift away from Omicron of a similar magnitude to the shift from Delta to Omicron); and (3) both severity increased to Delta and VFR increased to 10 (“increased severity and immune escape”). For all variant scenarios, we increase transmissibility by 10% to represent a variant that could replace Omicron.

### Forward simulations

For each scenario, we repeat the model simulation across 50 random seeds, with a simulation population size of 1 million, and summarise the median and 5–95% interval from the outputs. We use these outputs to calculate the daily infections and hospitalisations from 1 February 2020 to 31 December 2024, as well as the total infections, hospitalisations, and deaths from 1 July 2022 to the end of the simulation window. Setting characteristics and vaccine rollout assumptions are in **Table S5**.

### Cost-effectiveness

We estimate the cost per hospitalisation and death averted by calculating the total number of additional vaccine doses delivered, relative to the “3 doses only” (HIC) or “no additional doses” (LMIC) scenarios, divided by the difference in hospitalisations or deaths, multiplied by three illustrative costs for the vaccine unit price (US $2, $20, or $50 per dose), and assuming no difference in price between the different vaccine types.^25^

## Results

Across all scenarios, provided Omicron remains dominant and without any gradual immune escape or the emergence of new variants, we project COVID-19 reaching endemicity towards end-2022 or early-2023 (**Figures 1, 2, S5–7**). Within the HIC settings modelled, we project fewer hospitalisations and deaths from mid-2022 in high transmission countries (**Figures 1C–E, S7C–E**) due to the higher population immunity driven by hybrid immunity (**Figures 1B, S7B**). In both settings, hospitalisations and deaths can be substantially reduced by continuing regular boosting at either 6- or 12-monthly intervals to the highest risk age groups but these strategies have less impact on infection incidence (**Figures 1E, S7E**). In Category 1 countries, with an illustrative unit cost of $20 per dose for mRNA-1273.214, for yearly boosting this translates to $4,200 and $7,500 per hospitalisation averted for boosting 75+ and 60+ populations, respectively, and $13,500 and $31,900 per death averted (**Table 1**, values rounded to nearest $100). These same strategies are slightly more cost-effective in Category 3 countries due to the reduced alternative protection from prior infection-induced immunity. If the whole population (10+ years) is regularly boosted, we predict a substantial impact on transmission despite incomplete vaccine protection against infection, resulting in a more pronounced wave-like endemicity driven by population-level immune boosting and decay, and a lower endemic level (**Figures 1C–D, S7C–D)**. However, if the aim is solely to protect against hospitalisation and death then we estimate that higher efficiency (events averted per vaccine dose) can be achieved by regular vaccination of those aged 60+ (**Figures 1F, S7F**) with this reflected in the higher cost per hospitalisation or death averted of 10+ boosting (**Table 1**). The endemic prevalence – and hence the precise cost-effectiveness – is sensitive to our assumptions regarding the level of protection afforded by prior infection (**Figure S14**), with higher assumed levels of protective immunity from past infection resulting in lower endemic prevalence and therefore lower cost-effectiveness of vaccination.

**Table 1:** **Total additional infections, hospitalisations, and deaths averted, and total additional vaccine doses delivered for the Category 1 and 3 settings. We assume the mRNA-1273 is implemented for the first 2 doses and the first booster (dose 3), and the bivalent mRNA-1273.214 for subsequent booster doses.** Impact is expressed relative to the scenario where the primary series plus a booster is delivered to the 10+ years population, with no additional doses. Totals are shown for the period from 1 July 2022 to 31 December 2024. Unless otherwise specified, we assume no additional variant emergence beyond Omicron and its subtypes. The “new variant worse-case scenario” refers to a scenario where a new variant replaces Omicron over one month, starting 1 October 2023, with VFR = 10 relative to Delta, severity similar to Delta, and a transmission advantage of 10% relative to Omicron. The “variant drift scenario” refers to a scenario where the transmissibility and immune escape (VFR) gradually increase by 5% every 4 months, starting 1 April 2022. Values are the median estimate across 50 model simulations for each scenario. Total modelled events for each scenario are in **Table S6**.

| Vaccination scenario | Doses delivered per million population | Infections averted per thousand population | Hospitalisations averted per million population | Deaths averted per million population | Cost per hospitalisation averted (\$) | | | Cost per death averted (\$) | | |
| --- | --- | --- | --- | --- | --- | --- | --- | --- | --- | --- |
| Unit cost per vaccine dose (illustrative) |  |  |  |  | 2 | 20 | 50 | 2 | 20 | 50 |
| HIC with substantial prior transmission and high existing vaccine coverage |  |  |  |  |  |  |  |  |  |  |
| Boost 75+ yearly | 210015 | 73 | 1008 | 312 | 417 | 4167 | 10417 | 1346 | 13462 | 33656 |
| Boost 75+ 6-monthly | 350025 | 88 | 1134 | 350 | 617 | 6173 | 15433 | 2000 | 20001 | 50004 |
| Boost 60+ yearly | 675570 | 274 | 1796 | 424 | 752 | 7523 | 18808 | 3187 | 31867 | 79666 |
| Boost 60+ 6-monthly | 1125950 | 332 | 2116 | 491 | 1064 | 10642 | 26606 | 4586 | 45864 | 114659 |
| Boost 10+ yearly | 1337184 | 829 | 2496 | 524 | 1071 | 10715 | 26787 | 5104 | 51038 | 127594 |
| Boost 60+ yearly, new variant worst-case scenario | 675570 | 299 | 4799 | 1480 | 283 | 2827 | 7068 | 913 | 9129 | 22823 |
| Boost 60+ yearly, variant drift scenario | 675570 | 346 | 2278 | 524 | 593 | 5931 | 14828 | 2579 | 25785 | 64463 |
| Boost 10+ yearly,<br>new variant worst-case scenario | 1337184 | 831 | 5942 | 1724 | 450 | 4501 | 11252 | 1551 | 15513 | 38781 |
| Boost 10+ yearly,<br>variant drift scenario | 1337184 | 942 | 2959 | 621 | 904 | 9038 | 22595 | 4307 | 43066 | 107664 |
| HIC with limited prior transmission and high existing vaccine coverage |  |  |  |  |  |  |  |  |  |  |
| Boost 75+ yearly | 210015 | 5 | 1177 | 381 | 357 | 3569 | 8922 | 1102 | 11024 | 27561 |
| Boost 75+ 6-monthly | 350025 | 55 | 1373 | 430 | 510 | 5099 | 12747 | 1628 | 16280 | 40701 |
| Boost 60+ yearly | 675570 | 249 | 2236 | 540 | 604 | 6043 | 15107 | 2502 | 25021 | 62553 |
| Boost 60+ 6-monthly | 1125950 | 296 | 2606 | 592 | 864 | 8641 | 21603 | 3804 | 38039 | 95097 |
| Boost 10+ yearly | 1337184 | 802 | 3026 | 651 | 884 | 8838 | 22095 | 4108 | 41081 | 102702 |
| Boost 60+ yearly,<br>new variant worst-case scenario | 675570 | 312 | 5427 | 1650 | 249 | 2490 | 6224 | 819 | 8189 | 20472 |
| Boost 60+ yearly,<br>variant drift scenario | 675570 | 341 | 2676 | 634 | 505 | 5049 | 12623 | 2131 | 21311 | 53278 |
| Boost 10+ yearly,<br>new variant worst-case scenario | 1337184 | 815 | 6433 | 1882 | 416 | 4157 | 10393 | 1421 | 14210 | 35526 |
| Boost 10+ yearly,<br>variant drift scenario | 1337184 | 1032 | 3658 | 732 | 731 | 7311 | 18278 | 3654 | 36535 | 91338 |

**Figure 1:**
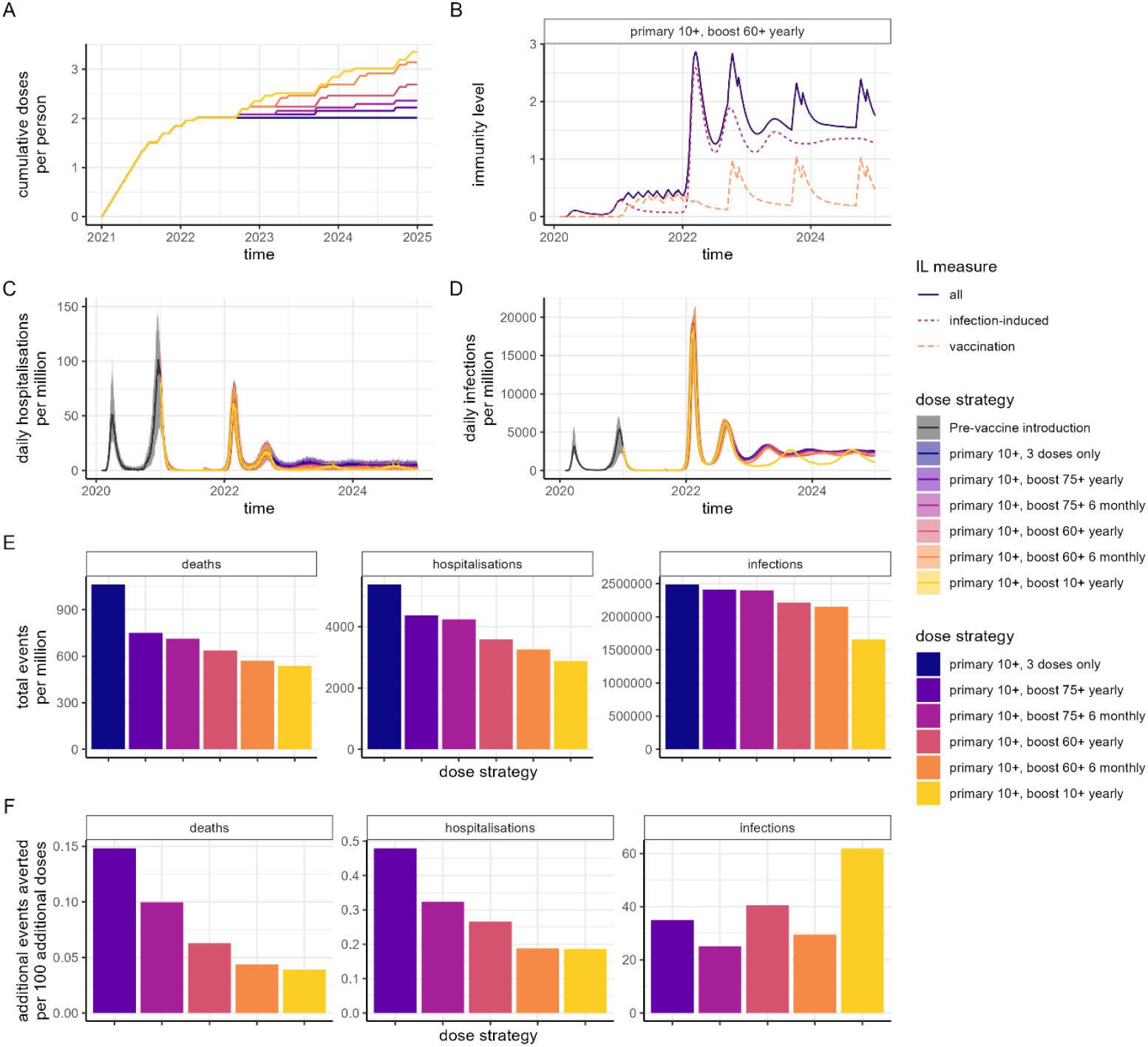
**Impact of vaccination in a high-income country setting with substantial prior transmission and high vaccine access. We assume mRNA-1273 is implemented for the first 2 doses and the first booster (dose 3), and the bivalent mRNA-1273.214 vaccine for subsequent booster doses. We assume no additional variant emergence beyond Omicron and its subtypes.** (A) Cumulative doses delivered per person over time, for a range of dose delivery strategies. In all strategies, the primary series was delivered to individuals 10 years and older, with scenarios of no additional doses; annual or 6-monthly boosters to the 75+ years population; annual or 6-monthly boosters to the 60+ years population; or annual boosters to the 10+ years population. (B) Mean infection-induced (pink dotted), vaccine-induced (orange dashed), and total (purple solid) immunity level (IL) over time for the “primary 10+, boost 60+ yearly” dose strategy. (C) Daily hospitalisations and (D) daily infections per million population for the six dose strategies, where the trajectory prior to vaccine introduction is shown in dark grey. (E) Total events (deaths, hospitalisations, and infection) per million population between 1 May 2022 and 31 December 2024 for each dose strategy. (F) Additional events averted per 100 additional doses over the same period relative to the “primary 10+, 3 doses only” dose strategy. Results for the scenario where the original mRNA-1273 vaccine continues to be administered, rather than switching to the bivalent product, are in **Figure S5**.

**Figure 2:**
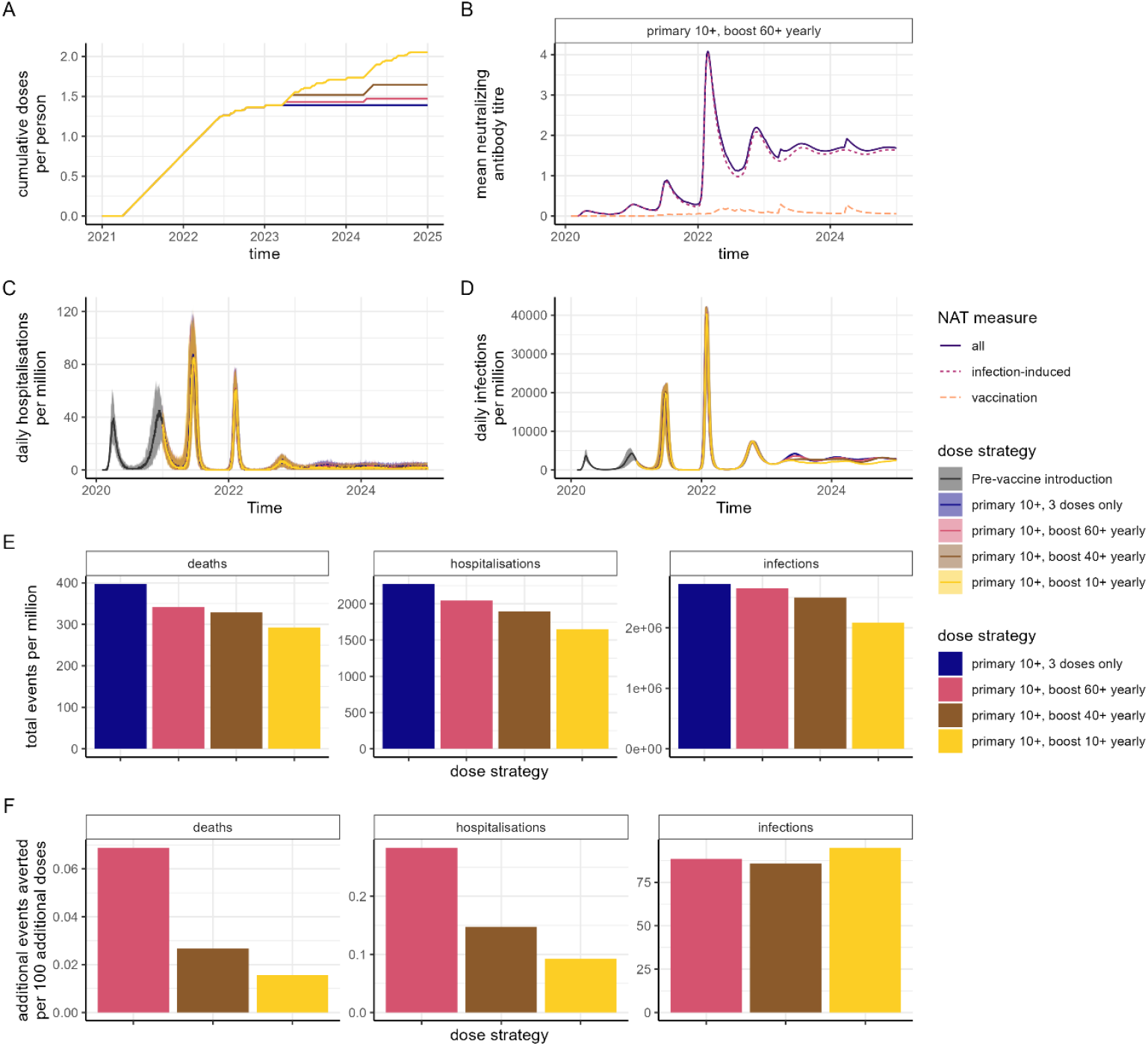
**Impact of vaccination in a lower-middle-income country setting with substantial prior transmission and moderate vaccine access. We assume AZD1222 is implemented for the first 2 doses, mRNA-1273 for the first booster (dose 3), and the mRNA-1273.214 bivalent vaccine for subsequent booster doses (doses 4 and 5). We assume no additional variant emergence beyond Omicron and its subtypes.** (A) Cumulative doses delivered per person over time, for a range of dose delivery strategies. In all strategies, the primary series was delivered to individuals 10 years and older, with scenarios of no additional doses; annual boosters to the 60+ years population; annual boosters to the 40+ years population; or annual boosters to the 10+ years population. (B) Mean infection-induced (pink dotted), vaccine-induced (orange dashed), and total (purple solid) immunity level (IL) over time for the “primary 10+, boost 60+ yearly” dose strategy. (C) Daily hospitalisations and (D) daily infections per million population for the dose strategies, where the trajectory prior to vaccine introduction is shown in dark grey. (E) Total events (deaths, hospitalisations, and infection) per million population between 1 July 2022 and 31 December 2024 for each dose strategy. (F) Additional events averted per 100 additional doses over the same time period relative to the “primary 10+, 3 doses only” dose strategy. Results for the scenario where the original mRNA-1273 vaccine continues to be administered, rather than switching to the bivalent product, are in **Figure S6**.

In LMIC populations that have experienced substantial prior transmission and have low vaccine coverage, we estimate that high infection-induced immunity is already present (**Figure 2B**). Assuming no further variants emerge, our projections suggest that these settings will reach endemic levels by 2023, although at a higher prevalence than in HIC settings where vaccination coverage is high (**Figure 2C–D**). Despite higher prevalence, hospitalisations and deaths are projected to be lower in LMIC compared to HIC settings due to a younger population combined with the broader protection generated from infection-induced immunity compared to vaccine-induced immunity. Hence, the costs per hospitalisation and death averted are substantially higher than in HICs (**Table 2**). At an illustrative unit cost of $2 per vaccine dose delivered (for mRNA-1273.214 but based on prior vaccination in these settings^25^) boosting the 40+ population would translate to $1,400 per hospitalisation averted and $7,400 per death averted. Total modelled doses, infections, hospitalisations and deaths for each Category and vaccination scenario are shown in **Tables S6–S9**, with the estimated impacts for the WHO vaccine coverage targets shown in **Figure S9** and **Table S10**.

**Table 2:** **Total additional infections, hospitalisations, and deaths averted, and total additional vaccine doses delivered for the Category 2 setting. We assume AZD1222 is implemented for the first 2 doses, mRNA-1273 for the first booster (dose 3), and the bivalent mRNA-1273.214 for subsequent booster doses (doses 4 and 5).** Impact is expressed relative to the scenario where the primary series plus a booster is delivered to the 10+ years population, with no additional doses. Totals are shown for the period from 1 July 2022 to 31 December 2024. Unless otherwise specified, we assume no additional variant emergence beyond Omicron and its subtypes. The “new variant worse-case scenario” refers to a scenario where a new variant replaces Omicron over one month, starting 1 October 2022, with VFR = 10 relative to Delta, severity similar to Delta, and a transmission advantage of 10% relative to Omicron. The “variant drift scenario” refers to a scenario where the transmissibility and immune escape (VFR) gradually increase by 5% every 4 months, starting 1 April 2022. Values are the median estimate across 50 model simulations for each scenario. Total modelled events for each scenario are in **Table S8**, with the total modelled events for the WHO coverage target scenario in **Table S10**.

| Vaccination scenario | Doses delivered per million population | Infections averted per thousand population | Hospitalisations averted per million population | Deaths averted per million population | Cost per hospitalisation averted (\$) | | | Cost per death averted (\$) | | |
| --- | --- | --- | --- | --- | --- | --- | --- | --- | --- | --- |
| Unit cost per vaccine dose (illustrative) |  |  |  |  | 2 | 20 | 50 | 2 | 20 | 50 |
| LMIC with substantial prior transmission and low existing vaccine coverage: default coverage target assumption |  |  |  |  |  |  |  |  |  |  |
| Boost 60+ yearly | 79918 | 71 | 226 | 55 | 707 | 7072 | 17681 | 2906 | 29061 | 72653 |
| Boost 40+ yearly | 256344 | 220 | 378 | 69 | 1356 | 13563 | 33908 | 7430 | 74303 | 185757 |
| Boost 10+ yearly | 672301 | 638 | 622 | 105 | 2162 | 21617 | 54043 | 12806 | 128057 | 320143 |
| Boost 60+ yearly, new variant worst-case scenario | 79918 | 70 | 525 | 200 | 304 | 3044 | 7611 | 799 | 7992 | 19980 |
| Boost 60+ yearly, variant drift scenario | 79918 | 78 | 248 | 58 | 644 | 6445 | 16112 | 2756 | 27558 | 68895 |
| Boost 10+ yearly, new variant worst-case scenario | 672301 | 529 | 1445 | 585 | 931 | 9305 | 23263 | 2298 | 22985 | 57462 |
| Boost 10+ yearly, variant drift scenario | 672301 | 614 | 563 | 98 | 2388 | 23883 | 59707 | 13720 | 137204 | 343011 |

We additionally find that in LMIC settings with high prior transmission, prioritising booster vaccinations in the highest-risk population has a slightly greater public health impact, reducing hospitalisations and deaths by ∼5–10%, compared to using these same doses to immunise younger age groups in an effort to reduce transmission (**Figure S8, Table S13**). We observed a larger difference in impact of these strategies for hospitalisations and deaths compared to infections (**Figure S8E**).

For the main results (described above) we simulated the estimated impact of the bivalent mRNA-1273.214 vaccine replacing the original vaccine from dose 4 onwards. Comparing these to corresponding scenarios in which the original mRNA-1273 product continues to be administered as booster doses, we estimate that switching to the bivalent vaccine product could avert around twice as many infections, hospitalisations, and deaths, and would reduce the cost per hospitalisation or death by around ∼50% over the time frame considered, assuming the cost per vaccine dose is equivalent (**Tables 1–2, Tables S11–S12, Figure S10**). However, continued administration of the original vaccines is anticipated to remain beneficial and efficient at the population level (**Figures S5–S6**).

**Figures 3 and S11–12** show the scenario where a gradual drift in the virus occurs, with such variants iteratively replacing the dominant circulating variant every 4 months with either 5% or 10% increased immune escape and transmissibility (referred to as “5% drift” and “10% drift”) and maintaining the same level of severity as for Omicron. Such a scenario mimics the patterns that have been observed in recent months with the global circulation of BA.4/BA.5 and more recent emergence in some areas of the XBB and BQ.1 variants. Here we observe a regular wave-like pattern driven by the changing variant properties resulting in increased infections and the subsequent increased population-level immunity. Administration of the bivalent vaccine is predicted to reduce more severe outcomes compared with continuing with the original vaccine, with a smaller impact on infections. However, over time this effect is reduced as further drift occurs and hence ongoing updates may be required.

**Figure 3:**
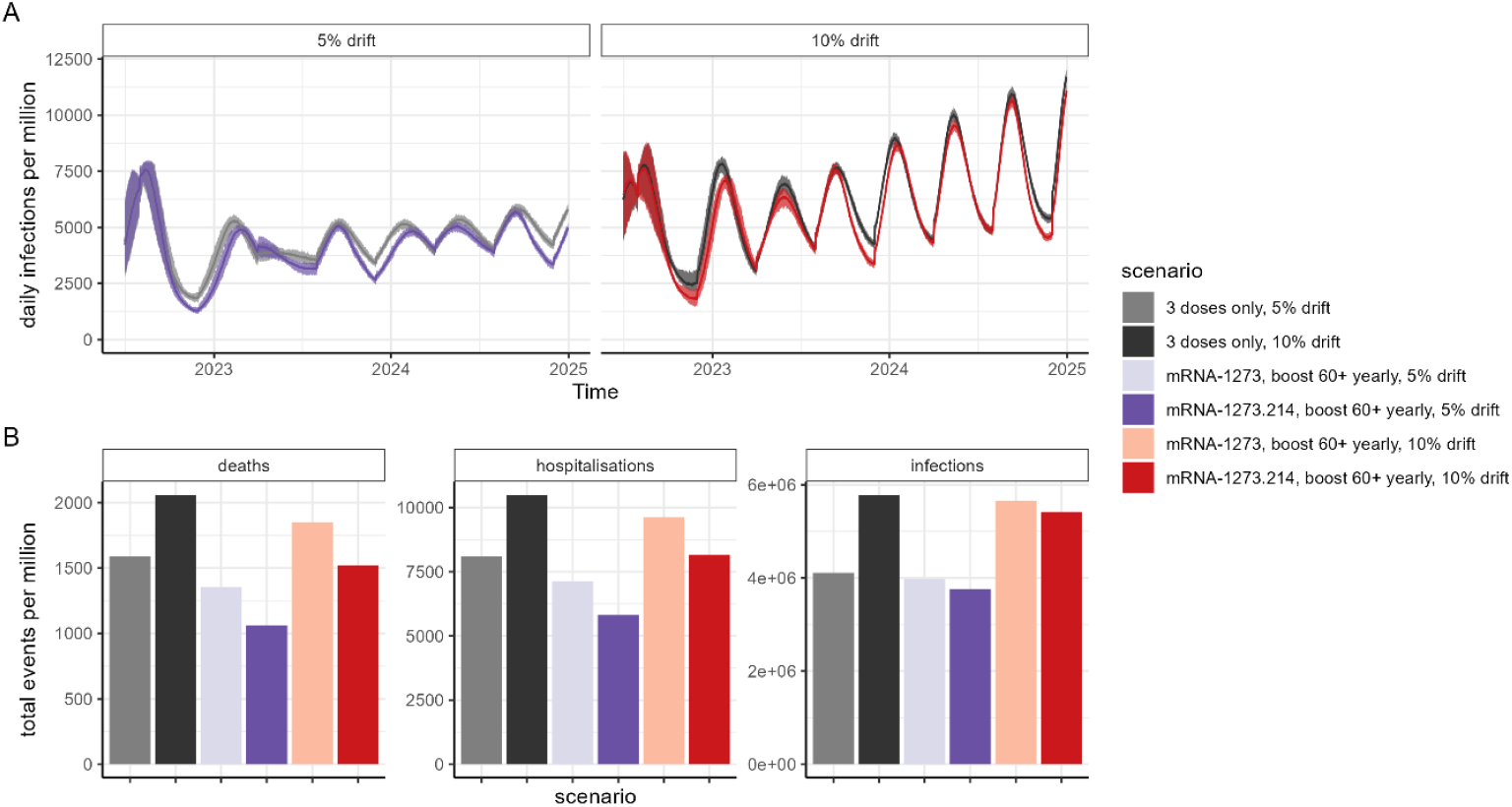
**Impact of vaccination in future scenarios where the virus continues to gradually evolve (or “drift”), in a high-income setting with substantial prior transmission (Category 1).** This is implemented in the model by increasing both the level of transmission and the VFR relative to Delta every 4 months, to represent both an increase in transmissibility and gradual immune escape. Here we show two levels of transmission and immune escape: 5% (purple/lilac) and 10% (red/orange). (A) Daily infections per million population assuming either 5% or 10% drift, either with the first three doses given only (light and dark grey), or with annual boosting to the 60+ population (purple and red). (B) Total events per million population between 1 July 2022 and end-2024, for the two “drift” scenarios, and for no boosting (light and dark grey) and boosting with either the original mRNA-1273 vaccine (lilac and orange), or with the bivalent mRNA-1273.214 vaccine from dose 4 onwards (purple and red). Results for the Category 2 and 3 settings are in **Figures S11–12**.

A completely new variant – simulated here as emerging in October 2023 – that replaces Omicron and its subtypes could rapidly result in a new epidemic wave, with the magnitude of this wave dependent on the properties of the variant (**Figure 4A–B, D–E**). Under a plausible worst-case scenario in which the variant is 10% more transmissible than Omicron, has a similar severity profile to Delta, and exhibits a shift in antigenic space similar to Omicron and therefore twice as far from the vaccines as was observed for Omicron, we predict levels of demand on health services similar to or exceeding those experienced during 2020 (**Figure 4C, F**). Under such a scenario, continued boosting scenarios become substantially more cost-effective despite the lower overall effectiveness of vaccination (**Tables 1–2**).

**Figure 4:**
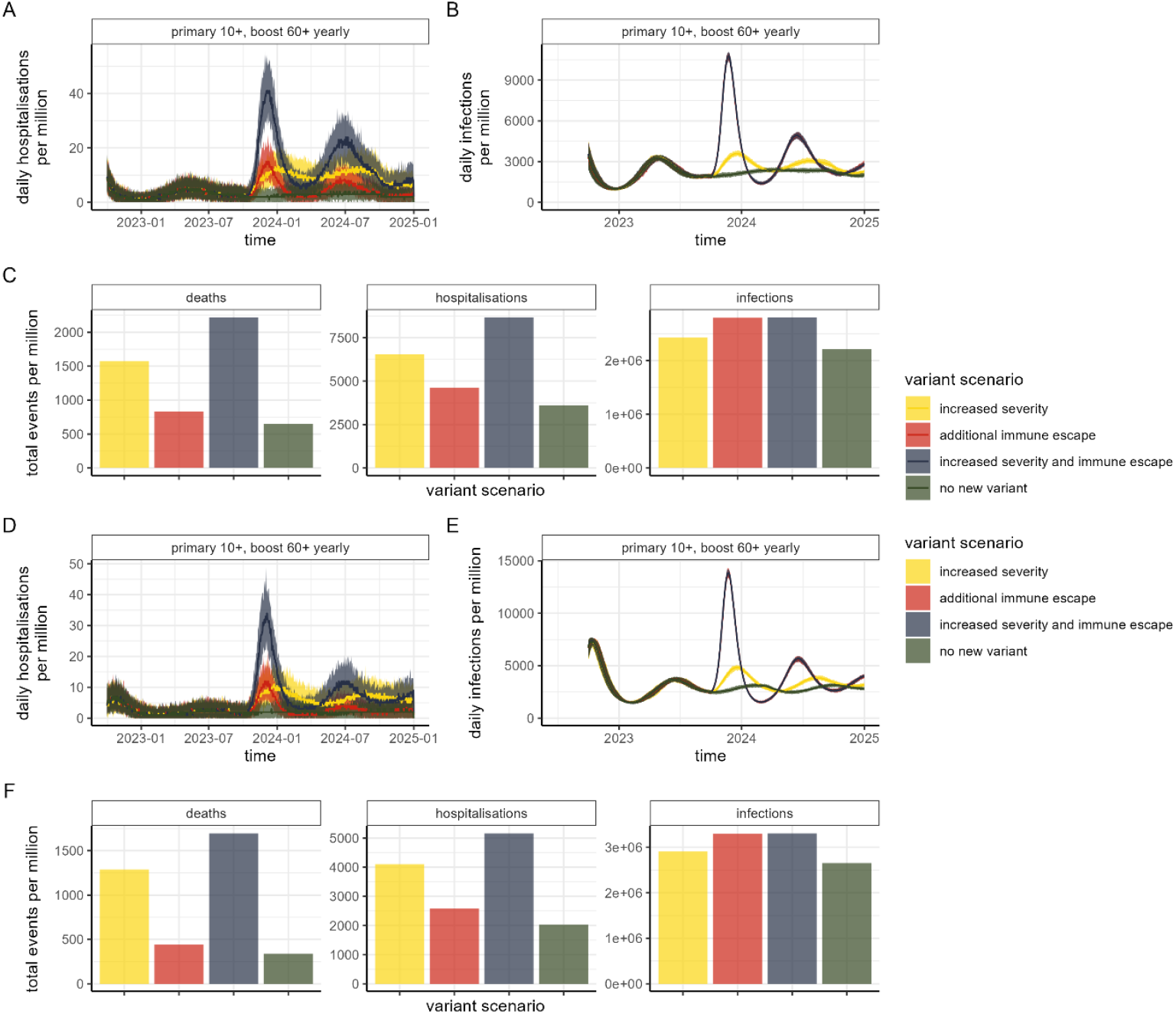
**Impact of vaccination in future scenarios where an additional variant of concern emerges from 1 October 2023. We assume the bivalent mRNA-1273.214 vaccine is implemented from dose 4.** Three variant scenarios are shown: increased severity, where the risk of hospitalisations and severe disease reverts to that of Delta (yellow); additional immune escape, where the VFR increases to 10 (red); and increased severity and immune escape, which assumes both Delta severity and a VFR of 10 (blue). This is compared to the scenario with no new variant (green). In all new variant scenarios, a 10% transmission increase is implemented from October 2023. (A) Daily hospitalisations, and (B) daily infections per million population for the high-income country setting with substantial prior transmission and high vaccine access (“Category 1”). (C) Total events (deaths, hospitalisations and infections) per million population for each variant scenario for the “Category 1” setting, between 1 July 2022 and end-2024. (D) Daily hospitalisations, and (E) daily infections per million population for the lower-middle-income country setting with substantial prior transmission and low vaccine access (“Category 2”). (F) Total events (deaths, hospitalisations and infections) per million population for each variant scenario for the “Category 2” setting, between 1 July 2022 and end-2024. Results for the Category 3 setting are in **Figure S13**.

## Discussion

Assuming continued circulation of the Omicron variant with or without a degree of virus drift, our projections suggest that most countries will move to an endemic level of SARS-CoV-2 circulation in the population towards the end of 2022 or beginning of 2023. The value of regular booster vaccination will therefore depend on assessment of the impact and cost-effectiveness of continued vaccination.

Our results demonstrate that the greatest impact on endemic prevalence can be achieved through regular boosting of the 10+ years population. However, the efficiency and cost-effectiveness of a boosting programme depends on the outcome measure; a strategy targeting only 75+ years averts the largest number of deaths and hospitalisations per dose, whereas a strategy targeting 10+ has the largest reduction in infections but is relatively inefficient in reducing severe outcomes. Similar patterns were obtained regardless of whether the country has previously experienced large waves of infection (and therefore has considerable infection-induced immunity) or whether the country had pursued a zero-COVID policy. However, in the zero-COVID policy setting or in LMICs with low vaccine coverage we generally estimate higher numbers of hospitalisations and deaths compared to settings with both high prior transmission and vaccine coverage. This is because either vaccine-induced immunity or infection-induced immunity alone is estimated to be less protective than the combination of vaccine- and infection-induced immunity (or hybrid immunity), as supported by immunogenicity studies. ^26,27^

Cost-effectiveness will likely be the metric driving future vaccine strategies. Our results demonstrate that across all settings considered, targeting the highest risk group is the most cost-effective strategy as judged by the cost per hospitalisation and death averted. With bivalent vaccines now being rolled out in some settings, we estimated that switching to such a variant-specific vaccine would reduce the cost per hospitalisation or death by around half. However, it should be noted that our estimates of bivalent vaccine effectiveness were based on immunogenicity studies and will therefore be sensitive to our fitted relationship between the underlying immunological mechanism and protection. To capture the full cost-effectiveness further information is needed on bivalent vaccine effectiveness and the comparative unit price of new products. We found that even continuing administration of the original vaccine products will reduce infections and severe outcomes in all settings. Furthermore, while estimating cost-effectiveness based on reductions in hospitalisations and deaths is relatively straightforward, such analyses do not account for the impact of high infection levels on long COVID incidence. COVID-19 hospitalisations and deaths in HICs have been concentrated in elderly populations; in contrast long COVID is reported across a wider age-range.^28,29^ Comprehensive cost-effectiveness analyses therefore need to consider the potential longer-term effects of this illness on quality of life and future productivity.

Our analysis is caveated by the uncertainty in the timing and impact of any new variant. By definition, any variant that can replace the currently circulating Omicron variant will either need to be more transmissible or exhibit significant immune escape. Given that antigenic mapping studies suggest that, to date, there is no clear pattern of antigenic drift,^30,31^ our assumptions should be regarded as plausible but illustrative rather than predictive. In addition, there is concern that a new variant could exhibit the increased severity seen with Delta. Our results illustrate that, under a worst-case scenario, an epidemic wave of similar magnitude to those experienced in the first year of the pandemic could occur, even with regular boosting to the highest risk age groups using bivalent vaccines. This ongoing uncertainty provides a further challenge in valuing vaccination programmes; whilst widespread boosting could mitigate the impact of a new variant and would be substantially more cost-effective if it did arise, such a boosting strategy is inefficient and therefore unlikely to be cost-effective if such a variant does not emerge. It will therefore be important for countries to consider other mitigation strategies such as timely provision of antivirals.

Our study has several limitations. First, the timing and magnitude of waves of SARS-CoV-2, the dominant circulating variant during these waves (particularly over the past 12 months), the timing and stringency of non-pharmaceutical interventions, and the vaccination response, has varied widely between countries. Our results are therefore illustrative and more detailed country-specific modelling will likely be required. Second, our immunological model is necessarily a simplification of the complex underlying immune response. The quality and durability of this response will likely vary by age; however, there are currently insufficient data to explore the impact of age on waning efficacy or immune escape from booster doses due to the shorter follow-up in younger populations. Furthermore, the degree to which prior immunity protects against future variants (including the currently circulating Omicron subtypes which are antigenically distinct from BA.1 estimates in the data used here)^30,31^ remains uncertain. Furthermore, the durability of infection-induced immunity compared to vaccine-induced immunity remains uncertain. Third, we only provide illustrative costing metrics as a first step towards broader cost-effectiveness analyses. Such analyses will depend on longer-term follow-up of the quality of life and persistence of disability following both mild infections and hospitalisation.

Our analyses demonstrate the importance of continued booster doses as part of the wider public health response to ongoing endemic transmission of SARS-CoV-2. Prioritising boosters to high-risk and older populations is an efficient strategy in terms of reducing hospitalisations and death, while managing finite healthcare resources; but further data are required to understand the cost-effectiveness of vaccinating a wider age group to protect against the consequences of long COVID.

## Data Availability

The model code is open access at https://mrc-ide.github.io/safir. All analysis code is available at https://github.com/mrc-ide/covid_booster_strategies.

## Acknowledgements

We thank Bob Verity, Nick Grassly, Sarah Pallas and the WHO SAGE working group on COVID vaccines for helpful comments and suggestions on earlier versions of this work.

## Funding

This work was supported by a grant from WHO. ABH acknowledges support from an NHMRC Investigator Grant and Imperial College Research Fellowship. PW is supported by an Imperial College Research Fellowship. OJW is supported by a Schmidt Science Fellowship in partnership with the Rhodes Trust. GC and ACG acknowledge support from The Wellcome Trust. ABH, PD, PW, GC, GB, SLW, NMF and ACG acknowledge funding from the MRC Centre for Global Infectious Disease Analysis (reference MR/R015600/1), funded by the UK Medical Research Council (MRC) and part of the EDCTP2 programme supported by the European Union. This work was additionally supported by the NIHR Health Protection Unit for Modelling and Health Economics (NMF: [NIHR200908]); and philanthropic funding from Community Jameel (PD, NMF).

## Author contributions

Conceptualization and study design: ACG, ABH, SLW, OJW, GB, PW, DOM; Individual-based model development: SLW, GC, OJW, PW, ABH; Analysis: ABH, ACG, JT, PD, SLW, DOM, PW; Visualization: ABH, JT, ACG, PD, SLW, DOM; Writing – original draft: ACG, ABH, SLW, JT, EMR; Writing – review & editing: All authors.

## Competing interests

ACG is a non-renumerated member of a scientific advisory board for Moderna, has received consultancy funding from GSK for educational activities related to COVID-19 vaccination and is a member of the CEPI scientific advisory board. She has received grant funding from Gavi for COVID-19 related work. ABH, PW and ACG have previously received consultancy payments from WHO for COVID-19 related work. ABH provides COVID-19 modelling advice to the New South Wales Ministry of Health, Australia. ABH was previously engaged by Pfizer Inc to advise on modelling RSV vaccination strategies for which she received no financial compensation. EMR is a non-remunerated member of the UK Vaccines Network, the UKRI COVID-19 taskforce and the British Society for Immunology Covid-19 taskforce.

## Notes

### Author Declarations

Ethics approval is not required as this is a modelling study using data in the public domain.

## References

1 Watson OJ, Barnsley G, Toor J, Hogan AB, Winskill P, Ghani AC. Global impact of the first year of COVID-19 vaccination: a mathematical modelling study. Lancet Infect Dis 2022; 22: 1293–302.

2 Magen O, Waxman JG, Makov-Assif M, et al. Fourth Dose of BNT162b2 mRNA Covid-19 Vaccine in a Nationwide Setting. N Engl J Med 2022. DOI:10.1056/NEJMoa2201688.

3 Bar-On YM, Goldberg Y, Mandel M, et al. Protection by a Fourth Dose of BNT162b2 against Omicron in Israel. New England Journal of Medicine 2022; 386: 1712–20.

4 Xue L, Jing S, Zhang K, Milne R, Wang H. Infectivity versus fatality of SARS-CoV-2 mutations and influenza. International Journal of Infectious Diseases 2022. DOI:10.1016/j.ijid.2022.05.031.

5 Wagner CE, Saad-Roy CM, Grenfell BT. Modelling vaccination strategies for COVID-19. Nat Rev Immunol. 2022; 22: 139–41.

6 Saadi N, Chi YL, Ghosh S, et al. Models of COVID-19 vaccine prioritisation: a systematic literature search and narrative review. BMC Med. 2021; 19. DOI:10.1186/s12916-021-02190-3.

7 Bubar KM, Reinholt K, Kissler SM, et al. Model-informed COVID-19 vaccine prioritization strategies by age and serostatus. Science (1979) 2021. https://www.science.org.

8 Saad-Roy CM, Morris SE, Metcalf CJE, et al. Epidemiological and evolutionary considerations of SARS-CoV-2 vaccine dosing regimes. Science (1979) 2021; 372: 363–70.

9 Kelly SL, le Rutte EA, Richter M, Penny MA, Shattock AJ. COVID-19 Vaccine Booster Strategies in Light of Emerging Viral Variants: Frequency, Timing, and Target Groups. Infect Dis Ther 2022; 11: 2045–61.

10 le Rutte EA, Shattock AJ, Chitnis N, Kelly SL, Penny MA. Modelling the impact of Omicron and emerging variants on SARS-CoV-2 transmission and public health burden. Communications Medicine 2022; 2. DOI:10.1038/s43856-022-00154-z.

11 Barnard RC, Davies NG, Munday JD, et al. Modelling the medium-term dynamics of SARS-CoV-2 transmission in England in the Omicron era. Nat Commun 2022; 13. DOI:10.1038/s41467-022-32404-y.

12 Organization WH. Strategic preparedness, readiness and response plan to end the global COVID-19 emergency in 2022. 2022 https://www.who.int/publications/i/item/WHO-WHE-SPP-2022.1.

13 Khoury DS, Cromer D, Reynaldi A, et al. Neutralizing antibody levels are highly predictive of immune protection from symptomatic SARS-CoV-2 infection. Nat Med 2021; 27: 1205–11.

14 Hogan AB, Doohan P, Wu SL, et al. Estimating long-term vaccine effectiveness against SARS-CoV-2 variants: a model-based approach. medRxiv 2023. DOI:10.1101/2023.01.03.23284131.

15 Chalkias S, Harper C, Vrbicky K, et al. A Bivalent Omicron-Containing Booster Vaccine against Covid-19. New England Journal of Medicine 2022; 387: 1279–91.

16 Altarawneh HN, Chemaitelly H, Ayoub HH, et al. Protective Effect of Previous SARS-CoV-2 infection against Omicron BA.4 and BA.5 subvariants. New England Journal of Medicine 2022; 387: 1620–2.

17 Regev-Yochay G, Gonen T, Gilboa M, et al. Efficacy of a Fourth Dose of Covid-19 mRNA Vaccine against Omicron. New England Journal of Medicine 2022; 386: 1377–80.

18 Charles G, Wu S. individual: An R package for individual-based epidemiological models. J Open Source Softw 2021; 6: 3539.

19 Hogan AB, Winskill P, Watson OJ, et al. Within-country age-based prioritisation, global allocation, and public health impact of a vaccine against SARS-CoV-2: A mathematical modelling analysis. Vaccine 2021; 39: 2995–3006.

20 Walker PGT, Whittaker C, Watson OJ, et al. The impact of COVID-19 and strategies for mitigation and suppression in low-and middle-income countries. Science 2020; 369: 413–22.

21 World Health Organization. WHO Coronavirus (COVID-19) Dashboard. 2022. https://covid19.who.int/table (accessed July 18, 2022).

22 World Health Organization Regional Office for Europe. WHO/Europe COVID-19 Vaccine Programme Monitor. 2022. https://worldhealthorg.shinyapps.io/EURO_COVID-19_vaccine_monitor/ (accessed July 18, 2022).

23 World Health Organization Regional Office for Africa.Africa COVID-19 Vaccination Dashboard. 2022. https://app.powerbi.com/view?r=eyJrIjoiOTI0ZDlhZWEtMjUxMC00ZDhhLWFjOTYtYjZlMGYzOWI4NGIwIiwidCI6ImY2MTBjMGI3LWJkMjQtNGIzOS04MTBiLTNkYzI4MGFmYjU5MCIsImMiOjh9 (accessed July 18, 2022).

24 World Health Organization. Strategy to Achieve Global Covid-19 Vaccination by mid-2022. 2021 https://www.who.int/publications/m/item/strategy-to-achieve-global-covid-19-vaccination-by-mid-2022 (accessed July 7, 2022).

25 UNICEF. Costs and predicted financing gap to deliver COVID-19 vaccines in 133 low-and middle-income countries. 2022 https://www.unicef.org/documents/costs-and-predicted-financing-gap-deliver-covid-19-vaccines-133-low-and-middle-income (accessed Jan 4, 2023).

26 Wang Z, Muecksch F, Schaefer-Babajew D, et al. Naturally enhanced neutralizing breadth against SARS-CoV-2 one year after infection. Nature 2021; 595: 426–31.

27 Bates TA, Mcbride SK, Leier HC, et al. Vaccination before or after SARS-CoV-2 infection leads to robust humoral response and antibodies that effectively neutralize variants. Sci Immunol 2022; 7: 8014.

28 Taquet M, Dercon Q, Luciano S, Geddes JR, Husain M, Harrison PJ. Incidence, co-occurrence, and evolution of long-COVID features: A 6-month retrospective cohort study of 273,618 survivors of COVID-19. PLoS Med 2021; 18. DOI:10.1371/journal.pmed.1003773.

29 Sudre CH, Murray B, Varsavsky T, et al. Attributes and predictors of long COVID. Nat Med 2021; 27: 626–31.

30 Rössler A, Netzl A, Knabl L, et al. BA.2 omicron differs immunologically from both BA.1 omicron and pre-omicron variants. medRxiv 2022. DOI:10.1101/2022.05.10.22274906.

31 Mykytyn AZ, Rissmann M, Kok A, et al. Omicron BA.1 and BA.2 are antigenically distinct SARS-CoV-2 variants. bioRxiv 2022. DOI:10.1101/2022.02.23.481644.

